# A Phase II study to evaluate the safety and efficacy of prasinezumab in early Parkinson’s disease (PASADENA): rationale, design and baseline data

**DOI:** 10.1101/2021.04.09.21251911

**Authors:** Gennaro Pagano, Frank G Boess, Kirsten I Taylor, Benedicte Ricci, Brit Mollenhauer, Werner Poewe, Anne Boulay, Judith Anzures-Cabrera, Annamarie Vogt, Maddalena Marchesi, Anke Post, Tania Nikolcheva, Gene G Kinney, Wagner M Zago, Daniel K Ness, Hanno Svoboda, Markus Britschgi, Susanne Ostrowitzki, Tanya Simuni, Kenneth Marek, Martin Koller, Jeff Sevigny, Rachelle Doody, Paulo Fontoura, Daniel Umbricht, Azad Bonni, PASADENA Investigators, Prasinezumab Study Group

## Abstract

**Background:** Currently available treatments for Parkinson’s disease (PD) do not slow clinical progression nor target alpha-synuclein, the main pathology associated with the disease.

**Objective:** The study objective was to evaluate the efficacy and safety of prasinezumab, a humanized monoclonal antibody that binds aggregated alpha-synuclein, in individuals with early PD. The study rationale, design, and baseline characteristics of enrolled subjects are presented here.

**Methods:** The PASADENA study is a multicenter, randomized, double-blind, placebo-controlled treatment study. Individuals with early PD, recruited across the US and Europe, received monthly intravenous doses of prasinezumab (1500 mg or 4500 mg) or placebo for a 52-week period (Part 1), followed by a 52-week extension (Part 2) in which all participants received active treatment. Key inclusion criteria were: aged 40–80 years; Hoehn & Yahr (H&Y) Stage I or II; time from diagnosis ≤2 years; having bradykinesia plus one other cardinal sign of PD (e.g. resting tremor, rigidity); DAT-SPECT imaging consistent with PD; and either treatment naïve or on a stable monoamine oxidase B (MAO-B) inhibitor dose. Study design assumptions for sample size and study duration were built using a patient cohort from the Parkinson’s Progression Marker Initiative (PPMI). In this report, baseline characteristics are compared between the treatment-naïve and MAO-B inhibitor-treated PASADENA cohorts and between the PASADENA and PPMI populations.

**Results:** Of the 443 patients screened, 316 were enrolled into the PASADENA study between June 2017 and November 2018, with an average age of 59.9 years and 67.4% being male. The mean time from diagnosis at baseline was 10.11 months, with 75.3% in H&Y Stage II. Baseline motor and non-motor symptoms (assessed using Movement Disorder Society – Unified Parkinson’s Disease Rating Scale [MDS-UPDRS]) were similar in severity between the MAO-B inhibitor-treated and treatment-naïve PASADENA cohorts (MDS-UPDRS Total score [standard deviation (SD)]; 30.21 [11.96], 32.10 [13.20], respectively). The overall PASADENA population (63.6% treatment naïve and 36.4% on MAO-B inhibitor) also showed a similar severity in MDS-UPDRS scores (e.g. MDS-UPDRS Total score [SD]; 31.41 [12.78], 32.63 [13.04], respectively) to the PPMI cohort (all treatment naïve).

**Conclusions:** The PASADENA study population is suitable to investigate the potential of prasinezumab to slow disease progression in individuals with early PD.

**Trial Registration:** NCT03100149

## Introduction

There is a high medical need to develop long-lasting therapies that can affect the underlying cause of Parkinson’s disease (PD) and, therefore, slow disease progression (1-3). Currently available treatments have powerful symptomatic effects, particularly on motor symptoms, but they do not address the pathological processes underlying the disease and do not prevent or slow clinical decline (2, 3). With the progressive loss of dopaminergic and non-dopaminergic neurons and synapses, available therapies gradually become less effective at controlling PD motor symptoms (4-6). Individuals with PD will invariably develop motor complications and lose their autonomy, adversely affecting their quality of life and placing a significant burden on caregivers, family members and healthcare systems (7-9). Despite the high prevalence and impact of non-motor symptoms on the quality of life of individuals with PD, treatment options for these symptoms are limited (10). A therapy that targets the underlying cause of the disease has the potential to slow motor progression as well as address non-motor symptoms (11, 12).

*Postmortem* findings suggest that the loss of dopaminergic neurons is accompanied spatially and temporally by the progressive development of intraneuronal Lewy pathology, which is a neuropathological hallmark of PD in distinct brain regions (13-16). Lewy pathology is abnormally enriched in alpha-synuclein, a protein with key functions in neurons (17). Although the etiology of PD is yet to be elicited, the spatio-temporal association between Lewy pathology and neurodegeneration, together with evidence from *in vitro* and *in vivo* models, suggests that pathologically aggregated forms of alpha-synuclein may contribute to axonal and neuronal damage, formation of Lewy pathology and consequent neuronal loss and disease progression (17-21).

Preclinical findings in cellular and animal models also support the hypothesis that certain aggregated forms of alpha-synuclein may be taken up by neurons and may induce the formation of intracellular alpha-synuclein inclusions in PD (19, 22-24). The appearance of intraneuronal inclusions throughout the central and peripheral nervous systems may arise upon propagation of Lewy pathology from neuron to neuron in a concerted manner by extracellular transfer of aggregated alpha-synuclein (20, 25-27).

Clinical evidence also supports the hypothesis that alpha-synuclein is a key driver in the etiology of PD. For instance, both missense mutations (28) and increased production of alpha-synuclein due to duplication or triplication of the synuclein gene (*SNCA*) (29-31) cause early-onset autosomal dominant PD, with virtually 100% penetrance (32). Although the exact patho-physiological mechanism in these genetic causes remains unclear, aggregation of alpha-synuclein due to missense mutation or overexpression is supposed to drive disease onset and progression (33). Direct transfer of aggregated alpha-synuclein from neuron to neuron has not been directly observed in humans. However, embryonic dopaminergic neurons transplanted into the striatum of individuals with PD harbored inclusions reminiscent of Lewy pathology approximately a decade after initial grafting (34, 35) which, together with replicated observations in animal models (22, 23), suggests the possibility of intercellular propagation of Lewy pathology. In support of a caudo-rostral propagation of Lewy pathology, molecular imaging studies demonstrate damage or dysfunction of noradrenergic and serotonergic pathways prior to the dopaminergic pathways in prodromal idiopathic and *SNCA* genetic PD (36, 37).

The growing understanding of the role of alpha-synuclein in the development of Lewy pathology and the pathogenesis of PD support the rationale that targeting alpha-synuclein may have therapeutic potential (38). Preclinical *in vivo* models of alpha-synucleinopathy, such as transgenic mice overexpressing wild-type human alpha-synuclein or that develop pathology upon intracerebral injection of aggregated recombinant alpha-synuclein, are valuable when studying drug mechanisms targeting alpha-synuclein. These models may help identify the downstream mode of action of therapeutic compounds. Indeed, neuropathological and behavioral deterioration in various mouse models of alpha-synuclein pathology was shown to be ameliorated by treatment with monoclonal antibodies binding to aggregated alpha-synuclein (12, 39-43).

Prasinezumab (previously known as RO7046015/PRX002) is an investigational, humanized monoclonal immunoglobulin G1 antibody directed against an epitope in the carboxyl terminus of human alpha-synuclein (11, 39, 40, 44). It binds to human aggregated alpha-synuclein with a high affinity and avidity (11, 39, 40). Preclinical pharmacologic studies to evaluate efficacy and potency of the murine form of prasinezumab (9E4) were performed in two transgenic mouse lines featuring alpha-synuclein aggregation disorders: Line D and Line 61 mice. The mice were treated with weekly intraperitoneal administration of murine version 9E4 over 5–6 months and showed reduced neuronal and synaptic loss and a reduction in intraneuronal build-up of alpha-synuclein pathology (measured as alpha-synuclein inclusions in cortical and subcortical regions), reduction of gliosis, and an improvement in both cognitive and motor behaviors (12, 39-41).

Although blockade of cell-to-cell transmission of alpha-synuclein by extracellular neutralization of pathogenic species has been proposed as the main mechanism of action of prasinezumab, evidence also supports potential clearance of alpha-synuclein species *via* the lysosomal pathway (12). Together, these preclinical data support the therapeutic potential of prasinezumab in slowing the progression of PD.

In a Phase I single-ascending-dose study in healthy volunteers and a Phase I multiple-ascending-dose study in individuals with PD, prasinezumab was safe, able to penetrate the blood–brain barrier (measured in the cerebrospinal fluid), and showed robust peripheral binding to alpha-synuclein (11, 44). Peripheral binding, measured as the lowering of circulating, free (unbound) serum alpha-synuclein, occurred within 1 hour of administration of prasinezumab and was maintained for longer durations with higher doses of prasinezumab (11). Results also demonstrated a dose-dependent increase of prasinezumab in cerebrospinal fluid concentration, which was approximately 0.3% relative to serum across all dose groups (11). Prasinezumab has a high binding affinity/avidity to aggregated alpha-synuclein vs. monomeric alpha-synuclein and, together with the observed cerebrospinal fluid concentrations achieved, it is predicted that >90% of aggregated alpha-synuclein will be engaged in the brain of individuals with PD at doses ≥1500 mg (11). The 1500 mg and 4500 mg doses were selected as both were expected to saturate the target in a Phase II study in individuals with early PD.

Here we report the study rationale, design, and baseline patient characteristics of PASADENA, a Phase II clinical trial testing efficacy and safety of prasinezumab in individuals with early PD. The study design and assumptions for sample size and study duration were built, in part, using a patient cohort from the Parkinson’s Progression Markers Initiative (PPMI) study group. The PPMI is a landmark, global, observational clinical study of individuals with PD designed to comprehensively evaluate cohorts of significant interest using advanced imaging, biological sampling and clinical and behavioral assessments to identify biomarkers of PD progression (45). We compared the baseline characteristics of the treatment-naïve and the monoamine oxidase B (MAO-B) inhibitor-treated cohorts of the PASADENA population. We also compared the baseline characteristics of the total PASADENA population with the characteristics of a subset of individuals with early PD enrolled in the PPMI study, which was selected using similar inclusion criteria to the PASADENA study (46).

## Methods

### Study design

The **P**hase II study of **A**nti alpha-**S**ynuclein **A**ntibo**D**y in **E**arly Parki**N**son’s dise**A**se (PASADENA) to evaluate the safety and efficacy of prasinezumab (NCT03100149) is an multicenter, randomized, double-blind, placebo-controlled study across approximately 60 sites in the United States, France, Austria, Germany and Spain. The study was designed to evaluate the efficacy and safety of intravenous prasinezumab (received every 4 weeks) in participants with early-stage PD (Hoehn and Yahr [H&Y] Stages I–II, time since diagnosis ≤2 years).

The study consists of two parts: a 52-week, double-blind, placebo-controlled treatment period (Part 1), followed by a 52-week extension period during which all participants received active treatment but remained blinded to original dose allocation (Part 2) (**Figure 1**). A 12-week safety follow-up was mandatory for all participants, regardless of whether cessation of treatment occurred after Part 1 or Part 2.

**Figure 1.**
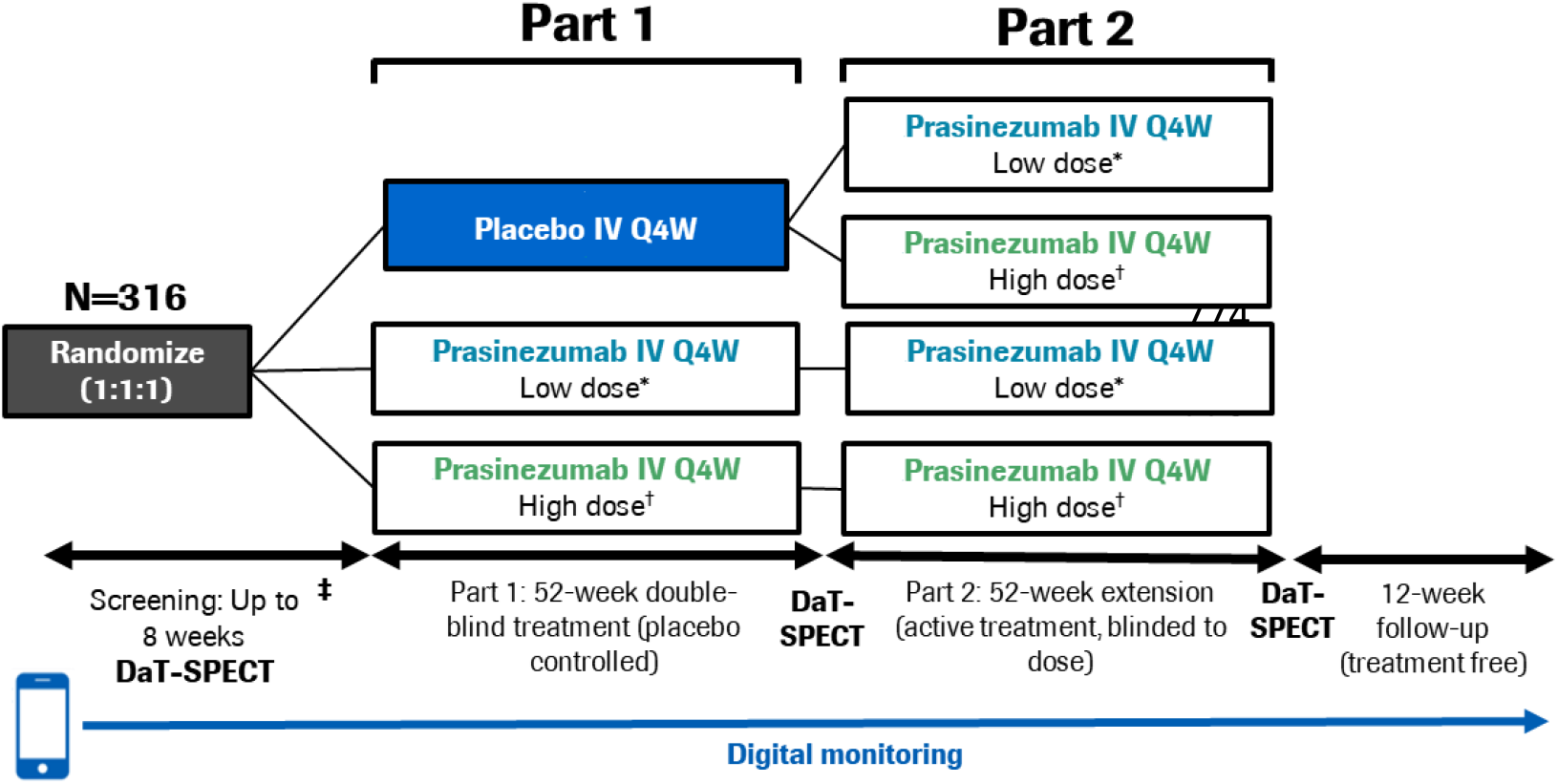
PASADENA study design schematic. *Low dose = 1,500 mg, ^†^High dose = 4,500 mg for ≥65 kg; 3,500 mg for <65 kg. DaT-SPECT, dopamine transporter imaging with single-photon emission computerized tomography; IV, intravenous; Q4W, every month.

In Part 1, participants were randomized with a 1:1:1 allocation ratio to either placebo, a high dose (4500 mg for body weight ≥65 kg; 3500 mg for body weight <65 kg) or a low dose (1500 mg for all body weights) of prasinezumab. Body weight has an effect on clearance (and volume of distribution) such that exposure increases in patients with lower body weight. Therefore, the use of an approximately 25% lower dose in participants with lower body weight (<65 kg) is implemented at high doses where there is an increased risk for infusion-related reactions. To reduce the risk of infusion-related reactions, participants in the high-dose group received a 2000 mg intravenous infusion on Day 1 followed by an up-titration to the full dose on Day 28 which they then received every 4 weeks. In addition, the first three study treatment infusions (irrespective of treatment allocation) were prolonged to 2 hours and were preceded by pre-medication with non-sedating antihistamine and acetaminophen (11). Randomization was stratified by sex, age group (<60 years vs. ≥60 years) and use of MAO-B inhibitor at baseline (yes vs. no).

Participants from Part 1 were eligible to continue to Part 2 provided dopamine transporter single-photon emission computerized tomography (DaT-SPECT) and magnetic resonance imaging (MRI) scans had been completed at screening and Week 52, and participants had received at least 10 doses of study treatment (placebo or prasinezumab) during Part 1.

In Part 2, participants randomized to treatment with prasinezumab in Part 1 remained on the same dose for the duration of Part 2. Those participants initially randomized to placebo were re-randomized to either 1500 mg or 4500 mg prasinezumab using a 1:1 allocation ratio. Randomization was stratified by dopaminergic therapy since start of study (yes vs. no), sex, age group at start of study (<60 years vs. ≥60 years) and use of MAO-B inhibitor at baseline (yes vs. no).

If symptomatic treatment was initiated during Part 1, investigators were required to record the reason(s) and the type and dose of symptomatic PD treatment prescribed. Participants who initiated symptomatic PD treatment could then continue in the study, as per their regular scheduled study visits. For participants who started dopaminergic treatment (levodopa or dopamine agonist), the Movement Disorder Society – Unified PD Rating Scale (MDS-UPDRS) including Part IV (motor assessment while on dopaminergic treatment) and digital biomarker in-clinic assessments at subsequent visits were performed in an “Off” state, i.e. patients had not received levodopa since the previous evening (>8 hours prior). The MDS-UPDRS Part III (motor assessment) was repeated at least 1 hour after receiving levodopa in the clinic (while patients are in an “On” state), along with digital biomarker in-clinic assessments.

### Study population

The inclusion and exclusion criteria were developed to select an early PD population with a measurable and precitable rate of progression over a 1-year period.

Key inclusion criteria included: idiopathic PD with bradykinesia and one of the other cardinal signs of PD (resting tremor, rigidity) and no other known or suspected cause of PD; aged 40–80 years; a DaT-SPECT consistent with PD; body weight range of ≥45 kg to ≤110 kg and a body mass index of 18 to 34 kg/m^2^; and either treatment naïve or on a stable dose of a MAO-B inhibitor for at least 90 days.

Key exclusion criteria included: medical history indicating a Parkinson syndrome other than idiopathic PD; known carriers of certain familial PD genes (*Parkin, PINK1, DJ1*); Mini Mental State Examination (MMSE) ≤25; use of any of the following: catechol-O-methyl transferase inhibitors (entacapone, tolcapone), amantadine or anticholinergics, or dopaminergic medication (levodopa and both ergot and non-ergot [pramipexole, ropinirole, rotigotine] dopamine agonists) for more than a total of 60 days or within 60 days of baseline and prior participation in any prasinezumab study.

A full list of inclusion and exclusion criteria can be found in Table 1 and Table 2 in the Supplementary Materials.

**Table 1.** PASADENA; MAO-B inhibitor-treated vs. treatment-naïve patients, and PASADENA (all patients) vs. PPMI. Demographics and baseline disease characteristics compared using the standardized mean difference

|  | PASADENA MAO-B inhibitor-treated patients (n=115) | PASADENA treatment-naïve patients (n=201) | SMD (CI) | PASADENA All patients (n=316) | PPMI PD patients (n=336) | SMD (CI) |
| --- | --- | --- | --- | --- | --- | --- |
| Age, years mean (SD) | 58.2 (9.00) | 60.8 (9.00) | -0.290*<br>(-0.520, -0.059) | 59.90 (9.10) | 61.30 (9.69) | 0.166<br>(0.012, 0.320) |
| Gender Male, n (%)† | 74 (64.3) | 139 (69.2) | -0.048<br>(-0.156, 0.060) | 213 (67.4) | 220 (65.5) | -0.019<br>(-0.092, 0.053) |
| Years of education, mean (SD) | 16.39 (5.20) | 15.22 (4.80) | 0.234 (0.006, 0.467) | 15.65 (4.99) | 15.53 (3.03) | -0.029<br>(-0.183, 0.125) |
| Time from diagnosis, months mean (SD) | 11.96 (6.10) | 9.06 (6.50) | 0.461*<br>(0.225, 0.689) | 10.11 (6.50) | 6.44 (6.30) | -0.573*<br>(-0.730, -0.417) |
| H&Y Stage II, n (%)† | 83 (72.2) | 155 (77.1) | -0.049<br>(-0.150, 0.051) | 238 (75.3) | 197 (59.0) | -0.167<br>(-0.238, -0.096) |
| RBDSQ (SD) | 3.51 (2.65) | 3.43 (2.75) | 0.031<br>(-0.199, 0.260) | 3.46 (2.71) | 4.14 (2.69) | 0.250<br>(0.095, 0.405) |
| DaT-SPECT contralateral putamen (SD) | 0.81 (0.24) | 0.78 (0.25) | -0.102<br>(-0.331, 0.128) | 0.80 (0.25) | 0.68 (0.27) | -0.445*<br>(-0.601, -0.289) |
| DaT-SPECT ipsilateral putamen (SD) | 1.02 (0.30) | 1.09 (0.33) | -0.194<br>(-0.421, 0.038) | 1.06 (0.32) | 0.96 (0.39) | -0.285*<br>(-0.439, -0.129) |
| MoCA (SD) | 28.27 (1.96) | 27.65 (2.04) | 0.309*<br>(0.076, 0.539) | 27.87 (2.03) | 27.24 (2.29) | -0.291*<br>(-0.446, -0.136) |
| SCOPA-AUT (SD) | 7.71 (4.82) | 8.25 (6.16) | -0.096<br>(-0.323, 0.136) | 8.05 (5.71) | 9.75 (6.23) | 0.284*<br>(0.128, 0.439) |
| MDS-UPDRS Total, mean (SD) | 30.21 (11.96) | 32.10 (13.20) | -0.150 (-0.378, 0.081) | 31.41 (12.78) | 32.63 (13.04) | 0.094<br>(-0.059, 0.248) |
| MDS-UPDRS Part I, mean (SD) | 4.49 (3.40) | 4.68 (4.06) | -0.051<br>(-0.279, 0.180) | 4.61 (3.83) | 5.60 (3.93) | 0.255*<br>(0.100, 0.409) |
| MDS-UPDRS Part IA, mean (SD) | 0.96 (1.54) | 1.27 (1.67) | -0.195<br>(-0.422, 0.037) | 1.16 (1.62) | 1.27 (1.57) | 0.071<br>(-0.083, 0.224) |
| MDS-UPDRS Part IB, mean (SD) | 3.53 (2.50) | 3.14 (2.93) | 0.045<br>(-0.185, 0.273) | 3.45 (2.78) | 4.33 (3.10) | 0.297*<br>(0.142, 0.451) |
| MDS-UPDRS Part II, mean (SD) | 5.19 (3.90) | 5.41 (4.13) | -0.055<br>(-0.284, 0.174) | 5.33 (4.04) | 6.12 (4.20) | 0.190<br>(0.036, 0.344) |
| MDS-UPDRS Part III, mean (SD) | 20.53 (8.81) | 22.01 (9.09) | -0.165<br>(-0.394, 0.065) | 21.47 (9.00) | 20.92 (8.88) | -0.062<br>(-0.216, 0.092) |
| MDS-UPDRS Part III Axial Symptoms, mean (SD)† | 0.77 (0.50) | 0.78 (0.57) | -0.004<br>(-0.233, 0.225) | 0.78 (0.54) | 0.68 (0.71) | -0.153<br>(-0.307, 0.001) |
| MDS-UPDRS Part III Bradykinesia, mean (SD)† | 9.91 (5.60) | 10.48 (5.49) | -0.102<br>(-0.331, 0.127) | 10.27 (5.53) | 10.60 (5.60) | 0.059<br>(-0.095, 0.212) |
| MDS-UPDRS Part III Rigidity, mean (SD)† | 3.90 (2.58) | 4.17 (2.83) | -0.103<br>(-0.331, 0.128) | 4.07 (2.74) | 3.86 (2.61) | -0.078<br>(-0.232, 0.075) |
| MDS-UPDRS Part III Resting Tremors, mean (SD)† | 2.71 (2.77) | 3.12 (2.74) | -0.147<br>(-0.377, 0.082) | 2.97 (2.75) | 2.58 (2.42) | -0.151<br>(-0.305, 0.003) |

\*Indicates not balanced covariates (>0.25 SMD). †For binary variables the table shows the difference in proportions. ‡Part III subscores are defined as: Bradykinesia (sum of item 3.4, finger tapping; item 3.5, hand movements; item 3.6, pronation-supination movements of hands; item 3.7, toe tapping; item 3.8, leg agility; item 3.9, arising from chair; item 3.13, posture; and item 3.14, body bradykinesia); Rigidity (sum of item 3.3, [Neck, Upper Limbs and Lower Limbs]); Resting tremors (sum of item 3.17, rest tremor amplitude [Lip/Jaw, Upper Limbs and Lower Limbs] and item 3.18, constancy of tremor); and axial symptoms (sum of item 3.10, gait; item 3.11, freezing of gait; and item 3.12, postural stability). CI, confidence interval; DaT-SPECT, dopamine transporter single-photon emission computerized tomography; H&Y, Hoehn and Yahr; MAO-B, monoamine oxidase B; MDS-UPDRS, Movement Disorders Society – Unified Parkinson's Disease Rating Scale; MoCA, Montreal Cognitive Assessment; PD, Parkinson's disease; PPMI, Parkinson's Progression Markers Initiative; RBDSQ, Rapid Eye Movement Sleep Behavior Disorder Screening Questionnaire; SCOPA-AUT, Scales for outcomes in Parkinson's disease autonomic dysfunction; SD, standard deviation; SMD, standardized mean difference.

### Objectives and endpoints

Clinical assessments performed at baseline and at different study visits are summarized in **Figure 2** and **Figure 3**. A full list of endpoints is available online in the PASADENA study protocol (NCT03100149) (47).

**Figure 2.**
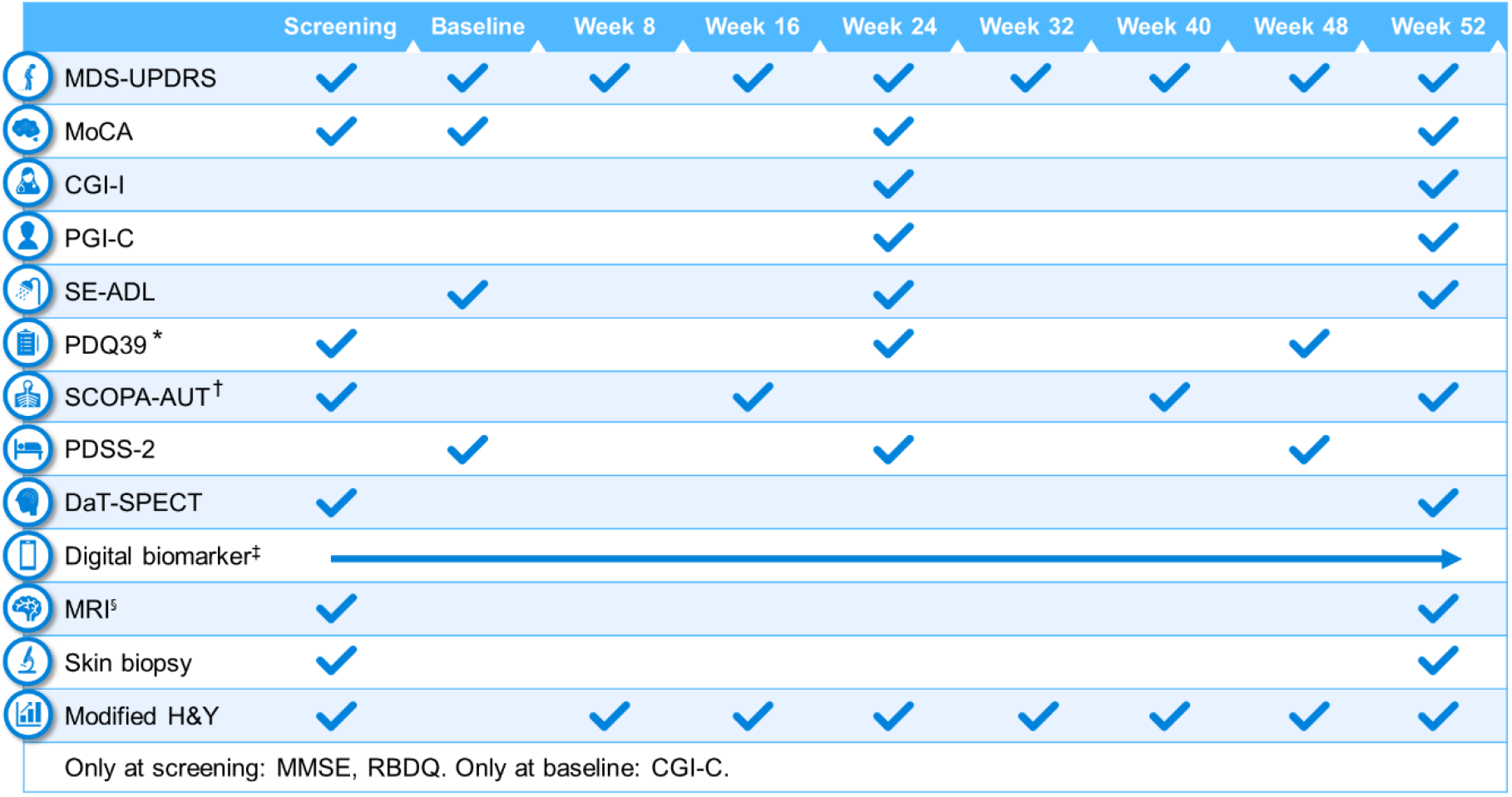
Schedule of activities in PASADENA Part 1. *Parkinson’s Disease Questionnaire – 39 (PDQ39) is at baseline, Week 20 and Week 48; ^†^SCOPA-AUT (Scales for outcomes in Parkinson’s disease autonomic dysfunction) is at baseline, Weeks 16, 28, 40 and 52. Digital includes PASADENA Digital Motor Score, Patient Global Impression of Severity, Daily diary, Patient Assessment of Constipation Symptoms (PAC-SYM), EuroQol-5D (EQ-5D) and Hospital Anxiety and Depression Scale (HADS). ^§^Magnetic resonance imaging (MRI) includes safety, diffusion tensor imaging (DTI), resting state and arterial spin labeling (ASL). CGI-I, Clinical Global Impression of Change; CGI-I, Clinical Global Impression of Improvement; DaT-SPECT, dopamine transporter imaging with single-photon emission computerized tomography; H&Y, Hoehn & Yahr; MDS-UPDRS, Movement Disorder Society – Unified Parkinson’s Disease Rating Scale; MMSE, Mini Mental State Examination; MoCA, Montreal Cognitive Assessment; PDSS-2, Parkinson’s Disease Sleep Scale Revised Version 2; PGI-C, Patient Global Impression of Change; RBDSQ, Rapid Eye Movement Sleep Behavior Disorder Screening Questionnaire; SE-ADL, Schwab and England Activities of Daily Living.

**Figure 3.**
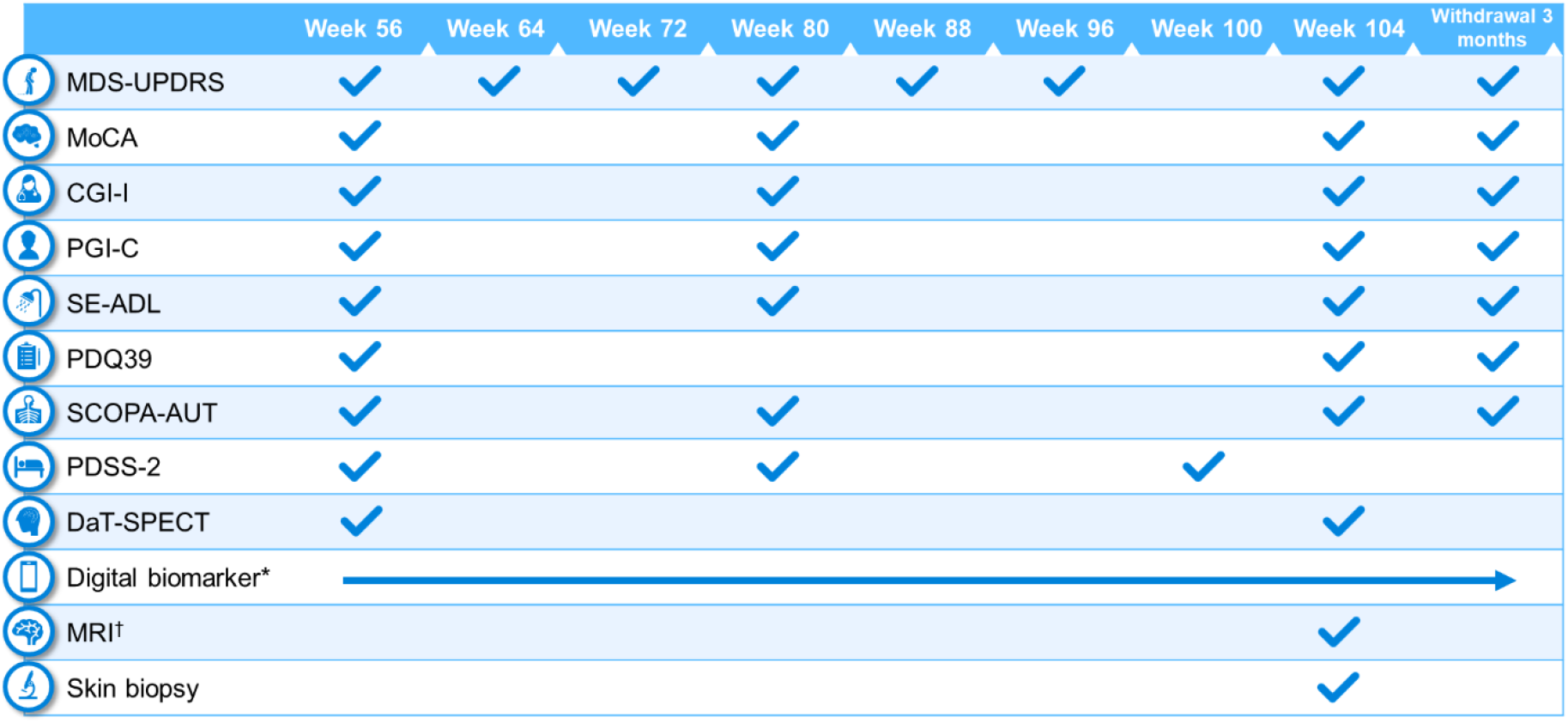
Schedule of activities in PASADENA Part 2. *Digital includes PASADENA Digital Motor Score, Patient Global Impression of Severity, Daily diary, Patient Assessment of Constipation Symptoms (PAC-SYM), EuroQol-5D (EQ-5D) and Hospital Anxiety and Depression Scale (HADS). ^†^Magnetic resonance imaging (MRI) includes safety, diffusion tensor imaging (DTI), resting state and arterial spin labeling (ASL). CGI-I, Clinical Global Impression of Improvement; DaT-SPECT, dopamine transporter imaging with single-photon emission computerized tomography; MDS-UPDRS, Movement Disorder Society – Unified Parkinson’s Disease Rating Scale; MoCA, Montreal Cognitive Assessment; PDQ-39, Parkinson’s Disease Questionnaire – 39; PDSS-2, Parkinson’s Disease Sleep Scale Revised Version 2; PGI-C, Patient Global Impression of Change; SCOPA-AUT, Scales for outcomes in Parkinson’s disease autonomic dysfunction; SE-ADL, Schwab and England Activities of Daily Living.

### Primary endpoint

The primary objective of the study was to assess the efficacy of prasinezumab 1500 mg and 4500 mg vs. placebo at Week 52 in enrolled participants. The primary endpoint was the change from baseline at Week 52 in MDS-UPDRS Total score (sum of Parts I, II and III) vs. placebo.

### Secondary endpoints

The effects of prasinezumab 1500 mg and 4500 mg vs. placebo at Week 52 on MDS-UPDRS Part IA, Part IB, Part I total, Part II total, Part III total and Part III subscores (bradykinesia, rigidity, resting tremor and axial symptoms) were included as secondary endpoints. Part I assessed non-motor experiences of daily living, with Part IA focused on complex behaviors (cognitive impairment, hallucinations and psychosis, etc.) and Part IB focused on non-motor experiences (sleep and urinary problems, constipation, pain, etc.). Part II assessed motor experiences of daily living (eating, dressing, handwriting, getting out of bed, etc.) and Part III assessed motor signs of PD (speech, finger tapping, bradykinesia, gait and freezing, etc.) (48). Part IA and Part III were administered by the study investigator, and Part IB and Part II were completed by the participant.

Other secondary endpoints included; Montreal Cognition Assessment (MoCA) Total score; Clinical Global Impression of Improvement (CGI-I); Patient Global Impression of Change (PGI-C); DaT-SPECT in the ipsilateral (to the clinically dominant side) putamen; Schwab and England Activity of Daily Living (SE-ADL) score; time to worsening in motor or non-motor symptoms (increase of ≥3 points in MDS-UPDRS Part I or MDS-UPDRS Part II); time to start of dopaminergic PD treatment (levodopa or dopamine agonists); and safety, tolerability, immunogenicity, and pharmacokinetics (PK) of prasinezumab. Safety and tolerability were assessed for up to 104 weeks, with or without dopaminergic treatment. PK of prasinezumab was assessed using population PK modeling.

### Exploratory endpoints

Exploratory endpoints of this study included: MDS-UPDRS Part III subscores determined by independent central raters (using video recordings to address consistency and accuracy in the trained site raters), imaging analysis of striatum, caudate and putamen (average, ipsilateral and contralateral) for DaT-SPECT binding ratio values, and the change from baseline on a sensor-based measure derived from Roche PD Mobile Application v2 digital biomarkers (49) (smartphone and wrist-worn wearable) assessments (see **Figure 4** for an overview of the remote monitoring tests). The analyses of primary and secondary endpoints were also repeated, with the results for the two prasinezumab 1500 mg and 4500 mg pooled doses vs. placebo, as a pre-specified exploratory analysis.

**Figure 4.**
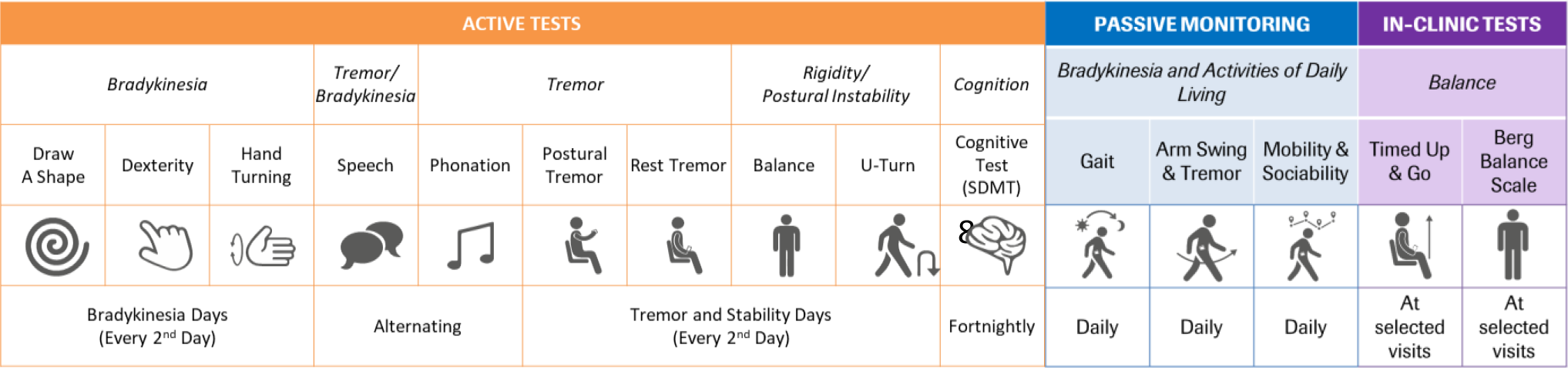
Table of digital measures included in the Roche Parkinson’s Disease Mobile Application v2. SDMT, Symbol Digit Modalities Test.

A full list of endpoints exploratory endpoints is included in Supplementary Table 3.

### PASADENA Digital Motor Score

Summary scores of sensor data from digital health technology tools should be developed independent of existing clinical data to ensure that the digital score does not inherit the shortcomings of the clinical measure (e.g. restriction of range, reduced resolution of scale). However, this requires independent longitudinal sensor datasets to build and validate such a digital score (49). Since such data are not yet available, a provisional single summary sensor-based measure, the ‘PASADENA Digital Motor Score’, reflecting global motor function was developed. Data from a PASADENA-like PPMI cohort were used to inform sensor feature selection for the PASADENA Digital Motor Score, as follows: MDS-UPDRS Part II and III item-level scores which significantly declined in Year 1 of PPMI were identified (n=21; whereby one item (posture) was not tested with the digital biomarker; final set of MDS-UPDRS items = 20). A blinded subset of PASADENA data (n=157) was used to map sensor feature data (aggregated over 2-week periods) onto each MDS-UPDRS item with the blinded PASADENA population. The set of sensor features was used to predict the sum of the 20 identified MDS-UPDRS items. PASADENA Digital Motor Scores were generated for every two weeks of the study and submitted to linear random coefficient (i.e. slope) models testing for differences in slopes between censored patients in each treatment group over the 26 two-week periods of the PASADENA study.

A second set of exploratory digital biomarker analyses comprised 17 individual pre-specified sensor features, which were selected based on a previous communication that reported cross-sectional correlations between sensor features and MDS-UPDRS scores (50) and available literature. These individual sensor features were also analyzed with linear random coefficient models testing for differences in slopes across treatment groups if the model’s residuals were normally distributed; if the residuals were not normally distributed, mixed effects models with repeated measures were applied.

### Standard protocol approvals, registrations, and patient consents

This study was conducted in full conformance with the International Conference on Harmonization E6 guideline for Good Clinical Practice and the principles of the Declaration of Helsinki, or the laws and regulations of the country in which the research was conducted, whichever afforded the greater protection to the individual. The study protocol, Informed Consent Forms, and any information given to the participant, were approved by the Institutional Review Board or Ethics Committee (NCT03100149).

### Sample size justification

A sample size of approximately 100 randomized participants per group (300 participants in total for the three groups) was estimated, which allowed for a power of approximately 80% at two-sided α-level of 20% to detect a three-point difference in MDS-UPDRS Total score between groups from baseline at Week 52. The power calculation was based on simulations of the mixed-effect model for repeated measures (MMRM) analysis planned for the primary efficacy variable. Assessments performed while on any symptomatic therapy started after randomization were not included in the analysis of the primary endpoint.

The assumptions on progression, variability, dropout rate and likelihood to start symptomatic therapy within the first 52 weeks of treatment, with or without a MAO-B inhibitor as background therapy, were derived from analyses based on the PPMI database and various sources of information from the literature (46, 51). The percentage of patients defined as non-evaluable at Week 52 was predicted to be 25% in the placebo group and 20% in the treatment groups. This estimate included non-evaluable data from patients who prematurely dropped out and/or started symptomatic therapy after randomization.

The sample size of 100 patients per arm also provided 76% power (α= 20%, two sided) to reject the null hypothesis, assuming a 37.5% reduction for the key secondary endpoint, the DaT-SPECT signal loss at Week 52 and the pairwise comparison of each active dose arm with placebo (46, 51).

### Covariate adjustment

Analyses of efficacy endpoints (primary, secondary, and exploratory) included the following covariates in the model: background therapy at baseline (MAO-B inhibitor treatment [yes vs. no]), age group (<60 years vs. ≥60 years), sex (male vs. female), DaT-SPECT binding ratio in the contralateral (to the clinically most affected side) putamen at baseline, and treatment group (4500 mg prasinezumab, 1500 mg prasinezumab or placebo). For each continuous endpoint the baseline of the endpoint variable was included in the model.

### Statistical analyses of primary efficacy endpoint

Change in MDS-UPDRS Total score from baseline vs. placebo was analyzed using an MMRM, with covariates described in the *Covariate adjustment* section as fixed effects. The model also included baseline MDS-UPDRS Total score, week of treatment (as a categorical factor), a treatment-by-week interaction term and an interaction term between baseline MDS-UPDRS by week. An unstructured variance-covariance matrix was used to model the random error. The model tested the null hypothesis of no treatment difference at a two-sided α-level of 20% for the following comparisons:

- 4500 mg or 3500 mg (high dose) prasinezumab vs. placebo
- 1500 mg (low dose) prasinezumab vs. placebo.

All the assessments flagged on and after the “first symptomatic PD treatment date” (either initiation of symptomatic PD treatment or a change to MAO-B inhibitor dose) were not included in the analysis. The primary endpoint of MDS-UPDRS Total score (sum of Parts I, II, and III) was re-analyzed with data from the two treatment arms pooled, and compared with placebo as an exploratory endpoint.

### Statistical analyses of secondary and exploratory efficacy endpoints

For the endpoints utilizing MDS-UPDRS Part IA, Part IB, Part I total, Part II total, Part III total, Part III subscores, CGI-I, and PGI-C, the information collected after symptomatic PD treatment was handled as for the primary analysis. The analysis of all other endpoints included all the data available regardless of start of symptomatic PD treatment.

An analysis of covariance (ANCOVA) was used to analyze the change from baseline at Week 52 in MoCA Total score and SE-ADL score with covariates described in the *Covariate adjustment* section. The change (between baseline and Week 52) in DaT-SPECT striatal binding ratio in the ipsilateral (to the clinically most affected side) putamen, was analyzed using ANCOVA. Arterial spin labeling (ASL) MRI was performed at sites with the technical capability. Effects are tested for in pre-specified regions of interest; striatum, caudate, and putamen (ipsilateral, contralateral, and average were assessed for each region). Between-group changes in ASL MRI over 1 year in regions of interest were also tested using ANCOVAs.

The CGI-I was intended as a measure of change in health status from baseline CGI - Severity of illness (CGI-S). For the CGI-I, patients were divided into one of two groups:

- ‘Responders’: Score of 1–4 (i.e. rated as “no change”, “minimally improved”, “much improved” or “very much improved”).
- ‘Progressors’: Score of 5–7 (i.e. rated as “minimally worse”, “much worse” or “very much worse”).

The proportion of patients rated by CGI-I grouping at Week 24 and Week 52 was analyzed using a logistic regression model. The estimated odds ratio for ‘responders’ and ‘progressors’ at Week 24 and Week 52 for treated patients compared with placebo were calculated with 80% confidence interval. Analysis of the PGI-C followed the same methodology outlined for the CGI-I above.

Time to worsening of motor or non-motor symptoms (of ≥3-point change from baseline in MDS-UPDRS Part I or Part II) and time to start of dopaminergic (levodopa or dopamine agonist) treatment were plotted using a Kaplan-Meier survival plot and analyzed using a Cox proportional hazards model to obtain a treatment difference between each of the prasinezumab dose levels against placebo. A 3-point minimum change in MDS-UPDRS Part II was chosen as this has been previously identified as the smallest change of score that is clinically meaningful to patients (52).

### Subgroup analyses

Subgroups with ≥20% of patients from the modified intent-to-treat population at baseline were analyzed. The model used for the primary endpoint was run in each subgroup, excluding the subgroup being analyzed if that was a covariate (e.g. MAO-B inhibitors at baseline [yes vs. no]).

The subgroup analyses were performed in the primary endpoint and the following secondary/exploratory endpoints:

- MDS-UPDRS Part I
- MDS-UPDRS Part II
- MDS-UPDRS Part III Total score and subscores
- MDS-UPDRS sums of Part II and III
- DaT-SPECT striatal binding ratio in the ipsilateral (to the clinically most affected side) putamen
- PASADENA Digital Motor Score
- MoCA score
- Composite time to event.

Subgroups included in analyses were:

- MAO-B inhibitors at baseline (yes vs. no)
- H&Y Stage at baseline (I vs. II)
- Rapid Eye Movement Sleep Behavior Disorder Screening Questionnaire (RBDSQ) at baseline (RBDSQ ≥5 vs. <5)
- Data-driven subphenotypes (Diffuse malignant vs. mild motor predominant vs. intermediate) at baseline
- Alpha-synuclein skin (positive vs. negative) (staining by immunohistochemistry on skin biopsy sections at baseline)
- DaT-SPECT striatal binding ratio in the ipsilateral (to the clinically most affected side) (very abnormal vs. abnormal) putamen.

For the derivations of the data-driven subphenotypes, scales were classified into: motor scales (MDS-UPDRS Part II and MDS-UPDRS-Part III) and non-motor scales (Scales for Outcomes in Parkinson’s Disease - Autonomic Dysfunction [SCOPA-AUT], RDBSQ and MoCA). After each one of the scales had been divided into percentiles, the data-driven subphenotypes were defined as follows:

- *Diffuse malignant*: Score on motor scales being greater than the 75^th^ percentile; and at least one score on a non-motor scale greater than the 75^th^ percentile; or all three non-motor scores greater than the 75^th^ percentile
- *Mild motor predominant*: Motor and all non-motor scores less than the 75^th^ percentile
- *Intermediate*: All those individuals not meeting criteria for other subtypes.

### Inclusion criteria for the PPMI cohort

The design of this study and the assumptions for progression, variability, dropout rate and likelihood to start symptomatic therapy within the first 52 weeks of treatment, with or without a MAO-B inhibitor, were derived from analyses of data collected in the PPMI observational clinical study. Details regarding the PPMI study have been previously published and are available at ppmi-info.org (53, 54). Patients from the PPMI population were selected for comparison with the PASADENA cohort using the following criteria, which align with the PASADENA study criteria: to have at least two of the following: resting tremor, bradykinesia, rigidity (must have either resting tremor or bradykinesia) and confirmation from imaging core that DaT-SPECT screening was consistent with dopamine transporter deficit. Data were downloaded in May 2020 and, from the total 423 individuals in the PPMI population, a cohort of 336 participants were selected based on the above criteria. No individuals in the PPMI cohort received MAO-B inhibitors at baseline.

### Statistical comparisons between PASADENA and PPMI cohorts

The standardized mean difference (SMD) in prognostic scores was used to assess differences between baseline data for the PASADENA and PPMI cohorts, and for the PASADENA MAO-B inhibitor-treated vs. treatment-naïve patient groups. The SMD was calculated as the absolute value in the difference in means of a covariate across the treatment groups, divided by the pooled SD. SMDs larger than 0.25 indicate that the groups were too different from one another for a reliable comparison of change from baseline in that variable (55). Overall, 20 covariates were selected for the analysis, including demographic, imaging and clinical assessment (MDS-UPDRS Part I, II and III) data.

## Results of baseline data analysis

### Baseline PASADENA demographics

A total of 443 patients were screened between June 2017 and November 2018 at 60 sites. Overall, 127 patients failed screening due to not meeting certain inclusion and exclusion criteria, such as brain DaT-SPECT screening consistent with PD, concomitant disease or condition within 6 months of screening and MMSE ≤25. Overall, 316 patients were enrolled at 57 centers across the following five countries: United States (160 patients [50.6%]), Spain (50 patients [15.8%]), France (65 patients [20.6%]), Germany (35 patients [11.1%]), and Austria (6 patients [1.9%]). Of those patients enrolled into the PASADENA study, 115 (36%) had received MAO-B inhibitor treatment at enrollment and 201 (64%) were treatment naïve. The mean (SD) age of PASADENA patients was 59.9 (9.10) years and the population included 213 (67.4%) men and 103 (32.6%) women, with a mean time from diagnosis of 10.11 (6.50) years and 238 (75.3%) individuals being H&Y Stage II. The mean (SD) MDS-UPDRS Total score at baseline was 31.41 (12.78) and the mean total scores for the individual parts were: Part I, 4.61 (3.83); Part II, 5.33 (4.04) and Part III, 21.47 (9.00). The mean baseline DaT-SPECT striatal binding ratios for the PASADENA population were 0.80 (0.25) for the contralateral putamen and 1.06 (0.32) for the ipsilateral putamen.

### MAO-B inhibitor treated vs. treatment-naïve patients in the PASADENA population

Within the PASADENA study population, patients treated with MAO-B inhibitors at baseline were on average younger (58.2 [9.00] years vs. 60.8 [9.00] years, respectively; SMD: –0.290) and had a longer time from diagnosis (11.96 [6.10] months vs. 9.06 [6.50] months, respectively; SMD: 0.461) vs. the treatment-naïve group. MAO-B inhibitor-treated patients also had a higher MoCA score vs. the treatment-naïve group (28.27 [1.96] vs. 27.65 [2.04], respectively; SMD: 0.309). All other baseline characteristics were balanced between patients who received a MAO-B inhibitor (n=115) and those who were treatment naïve (n=201) (**Table 1 and Supplementary Figure 1**).

### PASADENA population vs. PPMI cohort, selected using PASADENA eligibility criteria

When comparing the PASADENA population baseline characteristics with those of the PPMI cohort, MDS-UPDRS mean scores were lower for the PASADENA population for Part I (4.61 [3.83] vs. 5.60 [3.93], respectively; SMD: 0.255) and Part IB (3.45 [2.78] vs. 4.33 [3.10], respectively; SMD: 0.297) compared with the PPMI cohort. The PASADENA population had a longer average time from diagnosis (10.11 [6.50] years vs. 6.44 [6.31] years, respectively; SMD: –0.573) compared with the PPMI cohort. DaT-SPECT striatal binding ratios for the PASADENA population for the contralateral putamen (0.80 [0.25] vs. 0.68 [0.27], respectively; SMD: –0.445) and ipsilateral putamen (1.06 [0.32] vs. 0.96 [0.39], respectively; SMD: –0.285) to the clinically most affected side were both higher compared with the PPMI cohort. On average, PASADENA patients scored higher in MoCA (27.87 [2.03] vs. 27.24 [2.29], respectively; SMD: –0.291) and lower in SCOPA-AUT (8.05 [5.71] vs. 9.75 [6.23], respectively; SMD: 0.284) scores compared with the PPMI cohort. All other baseline characteristics were balanced between the PASADENA population and the PPMI cohort (**Table 1 and Supplementary Figure 2**).

## Discussion

PASADENA is the first Phase II study to test the efficacy of a monoclonal antibody binding aggregated alpha-synuclein to slow disease progression in early PD. The PASADENA study enrolled individuals diagnosed with early-stage PD, requiring that they be, *inter alia*, either treatment naïve or on stable treatment with MAO-B inhibitors and have a DaT-SPECT-confirmed dopaminergic deficit. In order for this Phase II proof-of-concept study to measure the disease-modifying potential of prasinezumab within the study period of 1 year, it was essential to define a population with a measurable rate of progression. The progression rate may depend on baseline disease severity and other factors (56). Several previous studies have reported that the progression rate in individuals with early PD is generally faster shortly after diagnosis of PD and before the start of levodopa or dopamine agonist therapy usage (46, 57). Individuals at this early stage of disease still have vulnerable dopaminergic neurons and protecting them against the development of further alpha-synuclein pathology may potentially slow motor disease progression. A 1-year treatment duration, if well powered, is expected to be sufficient to demonstrate relevant between-group differences, resulting from an effect of treatment on disease progression in individuals with early PD.

Current treatments for PD, such as levodopa, improve motor symptoms and aim to increase dopamine levels, compensating for the dopaminergic cell and synaptic loss (58). It is possible that the potential effects of treatment with a disease-modifying therapy on motor symptoms in individuals with early PD might be masked by these powerful symptomatic therapies (46, 54). People with early PD treated with MAO-B inhibitors have a reduced likelihood of starting levodopa or dopamine agonists therapy compared with placebo, while maintaining a relatively high progression rate (56, 57). Individuals with early PD who were either treatment naïve or treated with a MAO-B inhibitor at baseline were, therefore, included in Part 1 of the PASADENA study. The recruited PASADENA study population was also similar to other early PD therapeutic trial populations (56, 59, 60).

In this study, a comparison of the baseline characteristics of PASADENA study participants showed that those who received treatment with MAO-B inhibitors were younger with a longer time from diagnosis compared with the treatment-naïve group; however, the two groups had similar overall symptom severity at baseline. This observed similarity may be due to improvement in MDS-UPDRS scores as a result of MAO-B inhibitor treatment, which provides symptomatic relief to individuals with early PD by prolonging the action of dopamine in the brain (61, 62). The younger age may be due to the fact that individuals with older onset of PD usually receive levodopa rather than MAO-B inhibitors (63).

The design and assumptions of this study were informed from analyses of data collected in the PPMI observational clinical study. Therefore, it was important to compare the baseline characteristics of the PASADENA study population with the PPMI cohort to ensure the assumptions made were also valid for the PASADENA population. The PASADENA population and PPMI cohort showed different distributions for some demographic measures; for example, more PASADENA participants were in H&Y Stage II (75% vs. 59%) and had on average a 3.7-month-longer time from diagnosis than the PPMI cohort. However, the DaT-SPECT striatal binding ratios for both the ipsilateral and contralateral putamen suggested that the PASADENA population had a slightly less advanced disease in terms of severity and potentially of progression.

Dose selection for the PASADENA trial was based on data from previous studies. In a Phase I multiple-ascending-dose study (NCT02157714), individuals with mild-to-moderate PD who received prasinezumab up to 60 mg/kg intravenously every 4 weeks reported no serious or severe adverse events (11). Rapid-dose and time-dependent mean reductions from baseline vs. placebo in free serum alpha-synuclein levels of up to 97% were reported in trial participants after a single infusion at the highest dose (P=0.002), with similar reductions after two additional infusions. Mean cerebrospinal fluid concentration of prasinezumab also increased with dose, to approximately 0.3% relative to the concentration in serum across all dose cohorts. Currently, assays to quantify engagement of prasinezumab with aggregated forms of alpha-synuclein *in vivo* are not available. Thus, the dose selection for the Phase II study was primarily based on human serum and cerebrospinal fluid pharmacokinetic data extrapolation, and the relationship of these data with histopathological and functional endpoints. The doses used in the PASADENA trial were selected to fall in the therapeutic exposure range predicted from preclinical efficacy models; the high prasinezumab dose (4500 mg for body weight ≥65kg; 3500 mg for body weight <65kg) was selected to match exposure at the 60 mg/kg dose in the multiple-ascending-dose study, and the 1500 mg prasinezumab dose to yield exposure levels above those effective on alpha-synuclein pathology in the mouse model, with sufficient separation between the two to enable exposure response analyses. Both doses selected were expected to bind >90% of pathological aggregated alpha-synuclein, as well as monomers, in the central nervous system, thus both could show signal of efficacy on disease progression.

The effect of treatment with prasinezumab on clinical progression rate was determined using the MDS-UPDRS and was supported with a panel of exploratory biomarkers assessing the potential effects on PD pathology and progression of neuronal damage. The MDS-UPDRS is comprised of four parts: Part I, Mentation, Behavior, and Mood; Part II, Activities of Daily Living; Part III, Motor Examination; and Part IV, Complications of Therapy (48, 64). Each parkinsonian sign or symptom is rated on a 5-point scale (ranging from 0 to 4), with higher scores indicating more severe impairment (64). The MDS-UPDRS demonstrates good reliability, validity and sensitivity to change over a range of measures of time from diagnosis and severity (64). Previous studies in individuals with early PD have demonstrated a linear increase in MDS-UPDRS of approximately 6–12 points per year following diagnosis and prior to initiating symptomatic treatment (46, 54, 56, 57). A positive effect on MDS-UPDRS scores may, therefore, indicate a potential effect on global PD progression of prasinezumab. It is important to note that the increase in MDS-UPDRS scores in the treatment-naïve population with early PD is derived from the Part III motor examination scores; this population exhibits decline to a far lesser extent in activities of daily living (MDS-UPDRS Part I) and motor problems in daily life (MDS-UPDRS Part II) (64).

A striatal dopamine transporter deficit on dopamine transporter imaging by DaT-SPECT currently represents the most established imaging marker in PD, reflecting neurodegeneration in key brain regions affected by alpha-synuclein pathology. Regardless of dopaminergic treatment, individuals with early PD show the fastest decrease of DaT-SPECT signal (i.e. loss of dopaminergic terminals) resulting in an inverse exponential decline in striatal DaT-SPECT uptake values (65, 66). Therefore, DaT-SPECT will be used as a secondary outcome measure in the PASADENA study to determine the disease-modifying potential of prasinezumab.

Smartphones and smartwatches are built with high-quality sensors that, together with novel software technologies, enable the remote, non-invasive, frequent and sensitive measurement and analysis of motor and in PD (67-69). Digital monitoring of motor symptoms using smartphones has been previously used in the Phase I study of prasinezumab in individuals with PD (NCT02157714). Study participants completed a daily battery of tests and carried the phone with them throughout the day for passive monitoring. The study revealed high adherence and a strong correlation between smartphone sensor data and clinical measures of motor signs and, notably, the detection of clinical manifestations that were not apparent at site visits (70). A second version of this digital biomarker approach was therefore implemented in the PASADENA study to maximize the probability of detecting a potential therapeutic effect of prasinezumab and potentially provide new insights into the functioning and behavior of individuals with PD.

A definitive diagnosis of PD can only be made *post mortem* as biomarker tools that detect alpha-synuclein in the brain *in vivo* are not currently available (71). However, pathological forms of alpha-synuclein have been detected in peripheral neurons present in skin biopsy samples from individuals with PD; the degree of peripheral nerve pathology detected in samples was found to correlate with disease severity (72, 73). Longitudinal skin biopsy sampling has been implemented in the PASADENA study for the direct and *in vivo* assessment of alpha-synuclein pathology, and its progression in response to treatment with prasinezumab. Clinical data and samples, including skin biopsies, will be collected from DaT-SPECT negative screen failure participants to determine whether the detection of alpha-synuclein skin pathology may be used as a sensitive, specific, and less invasive tool to diagnose PD.

## Conclusions

The PASADENA Phase II study was designed to assess the safety and tolerability and clinical effect on disease progression of prasinezumab in patients with early PD. This study will focus on the effect of treatment in early PD, as disease progression is measurable and predictable in this cohort and will not be masked by treatment with dopaminergic therapy. The primary outcome measure will be supported by clinical measures and imaging to investigate the potential physiological impact of treatment. In addition, novel digital biomarkers will be used to assess potentially subtle effects of treatment on motor function in individuals with PD.

## Supporting information

Supplementary Table 1. Inclusion criteria

## Data Availability

Qualified researchers may request access to individual patient-level data through the clinical study data request platform (https://vivli.org/). Further details on Roche's criteria for eligible studies are available here (http://vivli.org/members/ourmembers/). For further details on Roche's Global Policy on the Sharing of Clinical Information and how to request access to related clinical study documents, see here (https://www.roche.com/research_and_development/who_we_are_how_we_work/clinical_trials/our_commitment_to_data_sharing.htm). 
Data used in the preparation of this article were obtained from the Parkinson's Progression Markers Initiative (PPMI) database (https://www.ppmi-info.org/data). For up-to-date information on the study, visit ppmi-info.org.

https://www.ppmi-info.org/data

## Data availability

Qualified researchers may request access to individual patient-level data through the clinical study data request platform (https://vivli.org/). Further details on Roche’s criteria for eligible studies are available here (https://vivli.org/members/ourmembers/). For further details on Roche’s Global Policy on the Sharing of Clinical Information and how to request access to related clinical study documents, see here (https://www.roche.com/research_and_development/who_we_are_how_we_work/clinical_trials/our_commitment_to_data_sharing.htm). Data used in the preparation of this article were obtained from the Parkinson’s Progression Markers Initiative (PPMI) database (ppmi-info.org/data). For up-to-date information on the study, visit ppmi-info.org.

## Ethics statement

Participants were identified for potential recruitment using site-specific recruitment plans prior to consenting to take part in this study. Recruitment materials for participants had received Institutional Review Board or Ethics Committee approval prior to use. The following Institutional Review Boards ruled on ethics of the PASADENA study: Ethikkommission der Medizinischen Universität Innnsbruck, Innsbruck, Austria; Comité de Protection des Personnes (CPP) Ouest IV, Nantes, France; Ethikkommission der Universität Leipzig and Geschäftsstelle der Ethikkommission an der medizinischen Fakultät der Universität Leipzig, Leipzig, Germany; Ethikkommission der Fakultät für Medizin der Technischen Universität München, München, Germany; Ethikkommission der Universität Ulm (Oberer Eselsberg), Ulm, Germany; Landesamt für Gesundheit und Soziales Berlin and Geschäftsstelle der Ethik-Kommission des Landes Berlin, Berlin, Germany; Ethikkommission des FB Medizin der Philipps-Universität Marburg, Marburg, Germany; Ethikkommission an der Medizinischen Fakultät der Eberhard-Karls-Universität und am Universitätsklinikum Tübingen, Tübingen, Germany; Ethikkommission an der Med. Fakultät der HHU Düsseldorf, Düsseldorf, Germany; Ethikkommission der LÄK Hessen, Frankfurt, Germany; CEIm Hospital Universitari Vall d’Hebron, Barcelona, Spain; Copernicus Group Independent Review Board, Puyallup, Washington, USA; Western Institutional Review Board, Puyallup, Washington, USA; The University of Kansas Medical Center Human Research Protection Program, Kansas City, Kansas, USA; Oregon Health & Science University Independent Review Board, Portland, Oregon, USA; Northwestern University Institutional Review Board, Chicago, Illinois, USA; Spectrum Health Human Research Protection Program, Grand Rapids, Michigan, USA; The University of Vermont Committees on Human Subjects, Burlington, Vermont, USA; Beth Israel Deaconess Medical Center Committee on Clinical Investigations, New Procedures and New Forms of Therapy, Boston, Massachusetts, USA; Vanderbilt Human Research Protection Program Health, Boston, Massachusetts, USA; Vanderbilt Human Research Protection Program Health, Nashville, Tennessee, USA; University of Maryland, Baltimore Institutional Review Board, Baltimore, Maryland, USA; University of Southern California Institutional Review Board, Los Angeles, California, USA; Columbia University Medical Center Institutional Review Board, New York, New York, USA; University of Southern California San Francisco Institutional Review Board, San Francisco, California, USA; University of Pennsylvania Institutional Review Board, Philadelphia, Philadelphia, USA; HCA - HealthOne Institutional Review Board, Denver, Colorado, USA. All Institutional Review Boards gave ethical approval of the study.

## Funding statement

The PASADENA study was funded by F. Hoffmann-La Roche Ltd.

PPMI, a public-private partnership, is sponsored by the Michael J. Fox Foundation (MJFF) for Parkinson’s Research and is co-funded by MJFF, AbbVie, Allergan, Amathus Therapeutics, Avid Radiopharmaceuticals, Bial Biotech, Biogen Idec, BioLegend, Bristol-Myers Squibb, Calico, Celgene, Denali Therapeutics Inc., 4D Pharma Plc, Eli Lilly and Company, F. Hoffmann-La Roche Ltd, GE Healthcare, Genentech Inc, GlaxoSmithKline, Golub Capital BDC, Handl Therapeutics, Insitro, Janssen Neuroscience, Lundbeck, Merck,

Meso Scale, Neurocrine Biosciences, Pfizer, Piramal, Prevail Therapeutics, Sanofi Genzyme, Servier, Takeda, Teva Pharmaceutical Industries Ltd, UCB, Verily Life Sciences and Voyager Therapeutics. Industry partners contribute to PPMI through financial and in-kind donations and have a lead role in providing feedback on study parameters through the Partners Scientific Advisory Board (PSAB). Through close interaction with the study, the PSAB is positioned to inform the selection and review of potential progression markers that could be used in clinical testing.

## Acknowledgments

The authors thank all the subjects who participated in this study. The authors thank Sarah Child of MediTech Media for providing medical writing support and Megan Speakman of MediTech Media for medical editing assistance, which were funded by F. Hoffmann-La Roche, in accordance with Good Publication Practice (GGP3) guidelines (http://www.ismpp.org/gpp3).

## Author contributions

GP, FGB, KIT, BM, WP, GGK, WMZ, DKN, SO, KM, MK and JS designed the study. GP, FGB, KIT, BM, WP, AnB, AV, MM, AP, SO and TS were involved in data collection. GP, KIT, BR, JAC, AV and MM analyzed the data. All authors were involved in data interpretation. GP and AzB drafted the work. GP, FGB, KIT, BR, BM, WP, JAC, AP, TN, GGK, WMZ, HS, SO, TS, KM, MK, JS, RD, PF, DU, and AzB critically revised important intellectual content. All authors revised and gave input on the article. GP and AzB provided final approval of the manuscript.

## Notes

### Competing Interest Statement

Gennaro Pagano is a full-time employee of F. Hoffmann-La Roche. Frank G Boess is a full-time employee of F. Hoffmann-La Roche. Kirsten I Taylor is a a full-time employee and shareholder of F. Hoffmann-La Roche. Benedicte Ricci is a full-time employee of F. Hoffmann-La Roche. Brit Mollenhauer reports personal fees from Roche, and served on the advisory board for Roche during the conduct of the study. Werner Poewe reports personal fees from Roche Pharmaceuticals during the conduct of the study; personal fees from Affiris, BIAL, Biogen, Britannia, Lilly, Lundbeck, Neuroderm, Neurocrine, Roche, Takeda, UCB, STADA and Zambon, outside of the submitted work. Anne Boulay was an employee of F. Hoffmann-La Roche, during the study. Judith Anzures-Cabrera is a full-time employee and shareholder of F. Hoffmann-La Roche. Annamarie Vogt is a full-time employee of F. Hoffmann-La Roche. Maddalena Marchesi is a full-time employee of F. Hoffmann-La Roche. Anke Post was an employee of F. Hoffmann-La Roche from 1.1.2017-30.11.2019. Tania Nikolcheva is a full-time employee and shareholder of F. Hoffmann-La Roche. Gene G Kinney is an officer, director and shareholder of Prothena Biosciences Inc, received funding for the work from F. Hoffmann-La Roche, and has a patent for Monitoring Immunotherapy Of Lewy Body Disease From Constipation Symptoms licensed to F. Hoffmann-La Roche. Wagner M Zago is an employee, officer and shareholder of Prothena Biosciences Inc, received funding for the work from F. Hoffmann-La Roche, and has a patent for Treatment Of Parkinson's Disease licensed to F. Hoffmann-La Roche. Daniel K Ness has nothing to disclose. Hanno Svoboda is an employee and shareholder of F. Hoffmann-La Roche. Markus Britschgi is a full-time employee and shareholder of F. Hoffmann-La Roche. Susanne Ostrowitzki is an employee of F. Hoffmann-La Roche and stock owner in the company. Tanya Simuni reports grants from NINDS, MJFF, Parkinson's Foundation, grants from Biogen, Roche, Neuroderm, Sanofi, Sun Pharma, Abbvie, IMPAX, Prevail, other from Acadia, Accorda, Adamas, Allergan, Amneal, Aptinyx, Denali, General Electric (GE), Kyowa, Neuroderm, Neurocrine, Sanofi, Sinopia, Sunovion, Roche, Takeda, Voyager, US World Meds, during the conduct of the study. Kenneth Marek reports grants and personal fees from Michael J Fox Foundation, personal fees from Roche, Takeda, Sanofi, Biohaven, GEHC, Inhibikase, Invicro, Hemacure, Neuron23, LTI and alkahest, outside the submitted work. Martin Koller was an employee of Prothena Biosciences Inc, during the study and received stock options during this time. Jeffrey Sevigny was an employee of F. Hoffmann-La Roche, during the study. Rachelle Doody is a full-time employee of Roche/Genetech and holds stocks and stocks options in the company. Paulo Fontoura is a full-time employee and shareholder of F. Hoffmann-La Roche. Daniel Umbricht is a former employee and shareholder of F. Hoffmann-La Roche. Azad Bonni is a full-time employee and shareholder of F. Hoffmann-La Roche.

### Clinical Trial

NCT3100149

