## Supplementary Table 1. Inclusion criteria for "A Phase II study to evaluate the safety and efficacy of prasinezumab in early Parkinson’s disease (PASADENA): rationale, design and baseline data"

**Supplementary material**

**Table 1. Inclusion criteria**

Participants met the following criteria for study entry:

1. Idiopathic PD with bradykinesia plus one of the other cardinal signs of PD (resting tremor, rigidity), without any other known or suspected cause of PD, and untreated or treated with a monoamine oxidase B (MAO-B) inhibitor.

2. Male or female, 40 to 80 years of age, body weight range of ≥45 kg/99 lbs to ≤110 kg/242 lbs and a body mass index of 18 to 34 kg/m^2^.

3. A diagnosis of PD for 2 years or less at screening.

4. Hoehn and Yahr (H&Y) Stage I or II.

5. A brain dopamine transporter single-photon emission computerized tomography (DaT-SPECT) screening, consistent with PD (central reading).

6. Clinical status did not require dopaminergic PD medication and was not expected to require dopaminergic therapy within 52 weeks from baseline.

7. If being treated for PD, must have received a stable dose of MAO-B inhibitor (rasagiline or selegiline) for at least 90 days prior to baseline and not expected to change within 52 weeks.

8. Able and willing to provide written informed consent and to comply with the study protocol according to International Conference on Harmonisation and local regulations.

9. For women of childbearing potential: use of highly effective contraceptive methods (that result in a failure rate of <1% per year) during the treatment period and for at least 30 days (or longer if required by local regulations) after the last dose of study drug.

- A woman is considered to be of childbearing potential if she is post-menarcheal, has not reached a post-menopausal state (≥12 continuous months of amenorrhea with no identified cause other than menopause), and has not undergone surgical sterilization (removal of ovaries and/or uterus).
- Examples of highly effective contraceptive methods (with a failure rate of <1% per year) include bilateral tubal ligation; vasectomized partner; established, proper use of hormonal contraceptives that inhibit ovulation; hormone-releasing intrauterine devices, and copper intrauterine devices.
- The reliability of sexual abstinence should be evaluated in relation to the duration of the clinical trial and the preferred and usual lifestyle of the patient. Periodic abstinence (e.g. calendar, ovulation, symptothermal, or post-ovulation methods) and withdrawal are not acceptable methods of contraception.

10. For men – use of contraceptive measures as defined below:

- With female partners of childbearing potential or pregnant female partners, men must use a condom during the treatment period and for at least 30 days (or longer if required by local regulations) after the last dose of study drug to avoid exposing the embryo. Men must refrain from donating sperm during this same period. The female partners should use a contraception method with a failure rate of <1% per year during the treatment period and for at least 30 days (or longer if required by local regulations) after the last dose of study drug.
- The reliability of sexual abstinence should be evaluated in relation to the duration of

the clinical trial and the preferred and usual lifestyle of the patient. Periodic abstinence (e.g. calendar, ovulation, symptothermal, or post-ovulation methods) and withdrawal are not acceptable methods of contraception.

**Table 2. Exclusion criteria**

Participants who met any of the following criteria were excluded from study entry:

**Current or past medical history**

1. Medical history indicating a Parkinson syndrome other than idiopathic PD, including but not limited to, progressive supranuclear gaze palsy, multiple system atrophy, drug-induced parkinsonism, essential tremor, primary dystonia.

2. Known carriers of certain familial PD genes (*Parkin, PINK1, DJ1*). Note: *GBA, synuclein, LRRK2* mutation carriers were allowed.

3. History of PD-related freezing episodes or falls.

4. A diagnosis of a significant central nervous system disease other than PD (including but not limited to Huntington’s disease, normal pressure hydrocephalus, cerebrovascular disease including stroke, fronto-temporal dementia, Alzheimer’s disease); history of repeated head injury; history of epilepsy or seizure disorder other than febrile seizures as a child.

5. Mini Mental State Examination (MMSE) ≤25.

6. Residing in a nursing home or assisted care facility.

7. History of or brain magnetic resonance imaging (MRI) screening scan indicative of clinically significant abnormality including, but not limited to, prior hemorrhage or infarct >1 cm^3^, >3 lacunar infarcts.

8. Concomitant disease or condition within 6 months of screening; or as specified below, that could interfere with; or treatment of which might interfere with, the conduct of the study; or that would, in the opinion of the Investigator, pose an unacceptable risk to the participant in this study; or interfere with the participant’s ability to comply with study procedures or abide by study restrictions; or with the ability to interpret safety data, including, but not limited to:
a) Autoimmune disease (however, well-controlled conditions such as, but not limited to, quiescent rheumatoid arthritis, controlled Type 1 diabetes, or mild-to-moderate psoriasis not requiring systemic medications may be acceptable after discussion with Sponsor/Medical monitors).
b) A history of cancer within 5 years of baseline with the exception of fully excised non-melanoma skin cancers or non-metastatic prostate cancer that has been stable for at least 6 months.
c) Any active infectious disease at baseline.
d) Current, or history of, alcohol or drug abuse or other dependence (except nicotine

dependence) within 2 years before screening.

e) Any febrile illness within 1 week prior to first dose administration.
f) Any current psychiatric diagnosis according to Diagnostic and Statistical Manual of

Mental Disorders fifth edition (DSM-5), International Statistical Classification of

Diseases and Related Health Problems 10th Revision (ICD-10) or equivalent, that

may interfere with the participant’s ability to perform the study and all assessments

(e.g. major depression, mental retardation, schizophrenia, bipolar disorder, etc.).

Note: Mild depression, depressive mood or mild anxiety arising in the context of PD, are not exclusionary.

9. The following cardiovascular conditions:
a) Myocardial infarction within 12 months of baseline.
b) Known history or documentation of uncontrolled hypotension or bradycardia on more than one occasion within 3 months prior to baseline.
c) Known history or documentation of uncontrolled hypertension on more than one occasion within three months prior to baseline.
d) Resting pulse rate greater than 100 or less than 45 bpm.
e) Clinically significant cardiovascular disease including any of the following: unstable

angina, decompensated congestive heart failure, clinically significant arrhythmias or symptomatic orthostatic hypotension.

f) A corrected QT interval measurement >450 ms for males or >470ms for females at screening, or a family history of long QT syndrome.
g) Intermittent second or third degree atrioventricular (AV) heart block or AV dissociation is excluded (asymptomatic patients with first degree AV block may be included).

10. Clinically significant abnormalities in laboratory test results at the screening visit, including hepatic and renal panels, complete blood count, chemistry panel and urinalysis, including:
a) Total bilirubin, alanine aminotransferase (ALT) or aspartate aminotransferase (AST) 2 times the upper limit of normal (ULN).
b) Serum creatinine >1.5 times the ULN.
c) Hematocrit (Hct) less than 35% for males and less than 32% for females, or absolute neutrophil cell count of <1500/µL (with the exception of a documented history of chronic benign neutropenia), or platelet cell count of <120,000/µL; international normalized ratio (INR) >1.4 (in patients not on anticoagulants) or other coagulopathy.
d) A clinically significant abnormal thyroid-stimulating hormone (TSH) test.
e) A positive urine drug screen for a drug of abuse.
   - For participants treated with selegiline, the amphetamine drug abuse test should be based on the results from a urine assay by liquid chromatography-mass spectrometry which is able to differentiate “false-positive methamphetamine” from “true-positive methamphetamine”.
   - For participants treated with benzodiazepines: a positive urine drug screen for benzodiazepines is allowed, provided that the prescription has been stable for 90 days prior to baseline (refer to exclusion criterion 15).
f) Positive result for acute or chronic infectious hepatitis B (HBV; [i.e. HBsAg positive

test]), for hepatitis C (HCV), or HIV 1 or 2. Successfully treated HCV patients (undetectable HCV RNA) are eligible for enrollment. Participants who are immune

due to HBV natural infection or HBV vaccination are eligible.

g) For women of childbearing potential, a positive urine or blood pregnancy test.

11. Lactating women.

**Medications and treatments**

1. Prior treatment with dopaminergic medication (e.g. levodopa or a dopaminergic agonist) with no clinical treatment response or a clinical treatment response inconsistent with PD (e.g. absence of observable response to a sufficiently high dose

of levodopa [i.e. ≥ 600 mg/day]).

2. Use of any of the following: catechol-O-methyl transferase inhibitors (entacapone, tolcapone), amantadine or anticholinergics or dopaminergic medication (levodopa and both ergot and non-ergot [pramipexole, ropinirole, rotigotine] dopamine agonists) for more than a total of 60 days or within 60 days of baseline.

3. Anti-epileptic medication for non-seizure-related therapy which had not remained stable for at least 60 days prior to baseline.

4. Antidepressant or anxiolytic use that had not remained stable for at least 90 days

prior to baseline. The use of fluoxetine and fluvoxamine was not permitted. For patients treated with a MAO-B inhibitor and an antidepressant (except fluoxetine and fluvoxamine), a 6-month period of stable and tolerated dosing before baseline was required.

5. Use of any of the following medications within 90 days prior to baseline; antipsychotics (including clozapine and olanzapine), metoclopramide, alpha methyldopa, flunarizine, amoxapine, amphetamine derivatives, reserpine, bupropion, buspirone, cocaine, mazindol, methamphetamine, methylphenidate, norephedrine, phentermine, phenylpropanolamine, and modafinil.

6. Participated in an investigational drug, device, surgical, or stem-cell study in PD.

7. Any prior treatment with an investigational PD-related vaccine (including active immunization or passive immunotherapy with monoclonal antibodies).

8. Prior participation in any RO7046015 or PRX002 study.

9. Receipt of any non-PD investigational product or device, or participation in a non-PD drug research study within a period of 30 days (or 5 half-lives of the drug, whichever was longer) before baseline.

10. Receipt of any monoclonal antibody or investigational immunomodulator within

180 days (or 5 half-lives, whichever is longer) before baseline (e.g. monoclonal

antibodies, intravenous immunoglobulin [IVIG], interleukin 2 [IL-2], interleukin 12

[IL-12], interferon or immunosuppressive drugs).

11. Immunomodulating drugs within 30 days prior to baseline.

12. Allergy to any of the components of prasinezumab such as citrate, trehalose and polysorbate (Tween) 20 or a known hypersensitivity or an infusion-related reaction to the administration of any other monoclonal antibody.

**Procedural**

1. Any contraindications to obtaining a brain MRI (e.g. claustrophobia unresponsive to reassurance or low dose of an anxiolytic agent, tooth implants) and any contraindications to obtain a DaT-SPECT (i.e. known hypersensitivity to the active substance or to any of the excipients). Patients with a hypersensitivity to iodine may receive an alternative thyroid-blocking agent (e.g. potassium perchlorate or sodium perchlorate).

2. For participants consenting to provide optional cerebrospinal fluid samples by lumbar puncture: lumbar puncture will only be performed if the participant does not have any contraindication to undergoing a lumbar puncture including, but not limited to: International normalized ratio >1.4 or other coagulopathy, platelet cell count of <120,000/µL, infection at the desired lumbar puncture site, taking anti-coagulant medication within 90 days of baseline (Note: low-dose aspirin (acetylsalicylic acid is permitted), severe degenerative arthritis of the lumbar spine, suspected non-communicating hydrocephalus or intracranial mass, prior history of spinal mass or trauma is/are identified. Participants failing to meet these criteria can still participate in the study and all other study assessments (with the exception of lumbar puncture) as appropriate. Participants could be excluded from the study if the cerebrospinal fluid has >5 white blood cells/mm^3^ (according to local laboratory assessment). This should be discussed with the Medical Monitor (e.g. if there is evidence that the spinal tap was traumatic, the participant may still be considered for study eligibility).

3. For skin biopsy at the cervical paravertebral region:
a) Condition that either precludes the safe performance of the skin punch biopsy or may interfere with obtaining evaluable skin tissue biopsies, including any previous or active significant dermatological disease (e.g. previous biopsy with any of the following findings: inflammatory disease, scar tissue, psoriasis, keloid formation,

skin cancer). Participants failing to meet these criteria can still participate in the study and all other study assessments (with the exception of skin biopsy) as appropriate.

4. Donation of blood over 500 mL within 3 months prior to screening.

**Table 3. Exploratory endpoints**

1. MDS-UPDRS Part III sub-scores were videotaped and the videos were centrally scored by a vendor to address consistency and accuracy in the trained site raters.

2. Time to starting dopaminergic PD therapy (levodopa or dopamine agonist).

3. Evaluated the effect of prasinezumab vs. placebo over 52 weeks on change from baseline in:
   - Modified H&Y
   - Hospital Anxiety and Depression Scale (HADS)
   - Patient Assessment of Constipation Symptoms (PAC-SYM)
   - Scales for outcomes in Parkinson’s disease autonomic dysfunction (SCOPA-AUT)
   - Parkinson’s Disease Sleep Scale Revised Version 2 (PDSS-2)
   - 39-item Parkinson’s Disease Questionnaire (PDQ-39)
   - European Quality of Life Questionnaire 5-level version (EQ-5D-5L)
   - Serum and cerebrospinal fluid biomarkers related to PD
   - Alpha-synuclein pathology in peripheral nerves (skin biopsies)
   - DaT-SPECT binding ratio values for: striatum, caudate and putamen (average, ipsilateral and contralateral)
   - Diffusion tensor imaging MRI for mean diffusivity and fractional anisotropy
   - Arterial spin labeling (ASL) MRI for cerebral blood flow.

4. Time to start or change of co-medication for non-motor symptoms that may have been related to either PD or imaging biomarkers (DaT-SPECT and ASL MRI).

5. Parkinson-related effects on the loss of autonomic tone as measured by heart rate variability.

6. Composite score of Part 2 and Part 3 sub-items.

7. MDS-UPDRS Part IV at Week 52 in participants who started dopaminergic therapy (levodopa or dopamine agonist).

8. Change from baseline in following patient-reported outcomes (PRO) were assessed:
   - Diary questions
   - PAC-SYM questionnaire
   - HADS total score and sub-scales:
     - anxiety (HADS-A)
     - depression (HADS-D).
   - EQ-5D-5L questionnaire

9. Change from baseline over 52 weeks in following Roche PD Mobile Application v2 digital biomarker [40] (smartphone and wrist-worn wearable assessments) were assessed:

- Sensor data collected during “Active Tests”, assessing motor symptoms (upper and lower body movement, upper limb dexterity, voice/speech) and non-motor symptoms (including an electronic version of the Symbol Digital Modalities Test to measure attention and executive function).
  - Sensor data collected during “Passive Monitoring” assessing activity, movement and motor symptoms associated with routine daily living.
  - Sensor data collected during “In-Clinic Assessments”, including the Timed Up and Go Test and selected items from the Berg Balance Scale.

10. Since the two doses of prasinezumab were expected to reach occupancy of >90% of the target in central nervous system, pooling of patients in the two prasinezumab cohorts was pre-specified for a subset of exploratory endpoints.
