## Supplementary Figure 1 for "A Phase II study to evaluate the safety and efficacy of prasinezumab in early Parkinson’s disease (PASADENA): rationale, design and baseline data"

**Figure 1.** Covariate balance between the treatment-naïve and MAO-B inhibitor subgroups of the PASADENA population (Love plot)

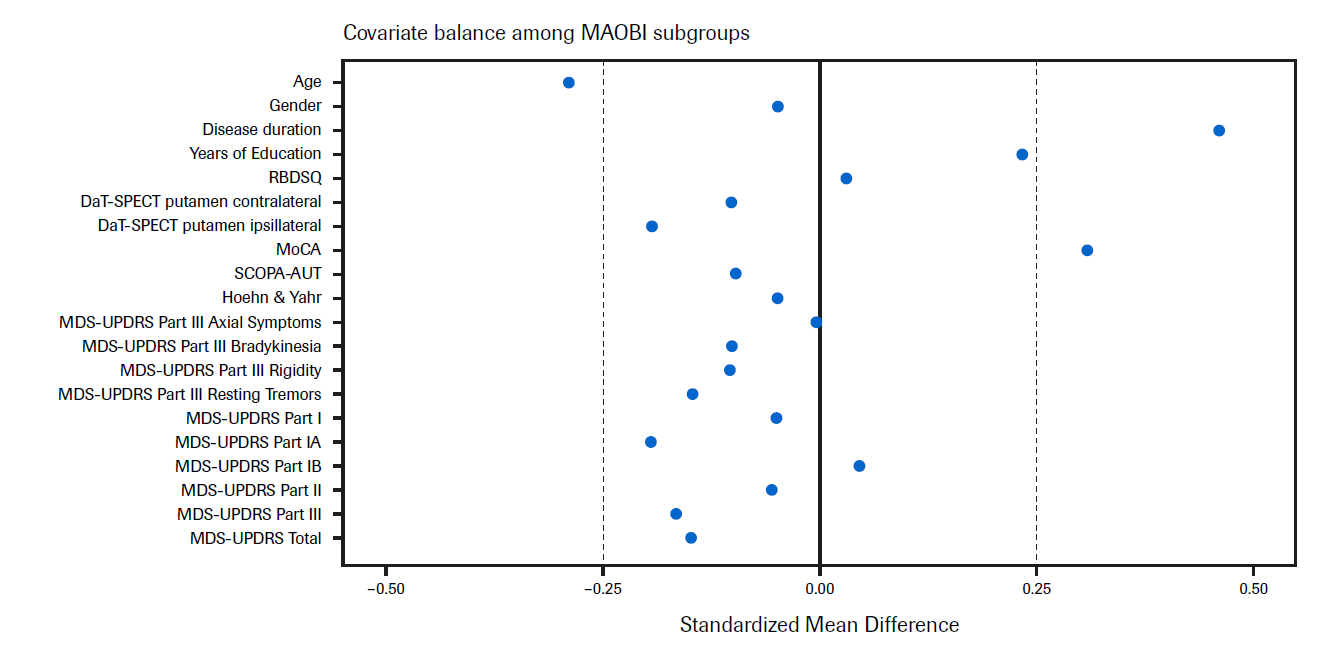

DaT-SPECT, dopamine transporter single-photon emission computerized tomography; MDS-UPDRS, Movement Disorder Society – Unified Parkinson’s Disease Rating Scale; MoCA, Montreal Cognitive Assessment; PPMI, Parkinson’s Progression Marker Initiative; RBDSQ, Rapid Eye Movement Sleep Behavior Disorder Screening Questionnaire; SCOPA-AUT, Scales for outcomes in Parkinson’s disease autonomic dysfunction.
