## Supplementary Figure 2 for "A Phase II study to evaluate the safety and efficacy of prasinezumab in early Parkinson’s disease (PASADENA): rationale, design and baseline data"

**Figure 2.** Covariate balance between the PASADENA population and the PPMI cohort (Love plot)

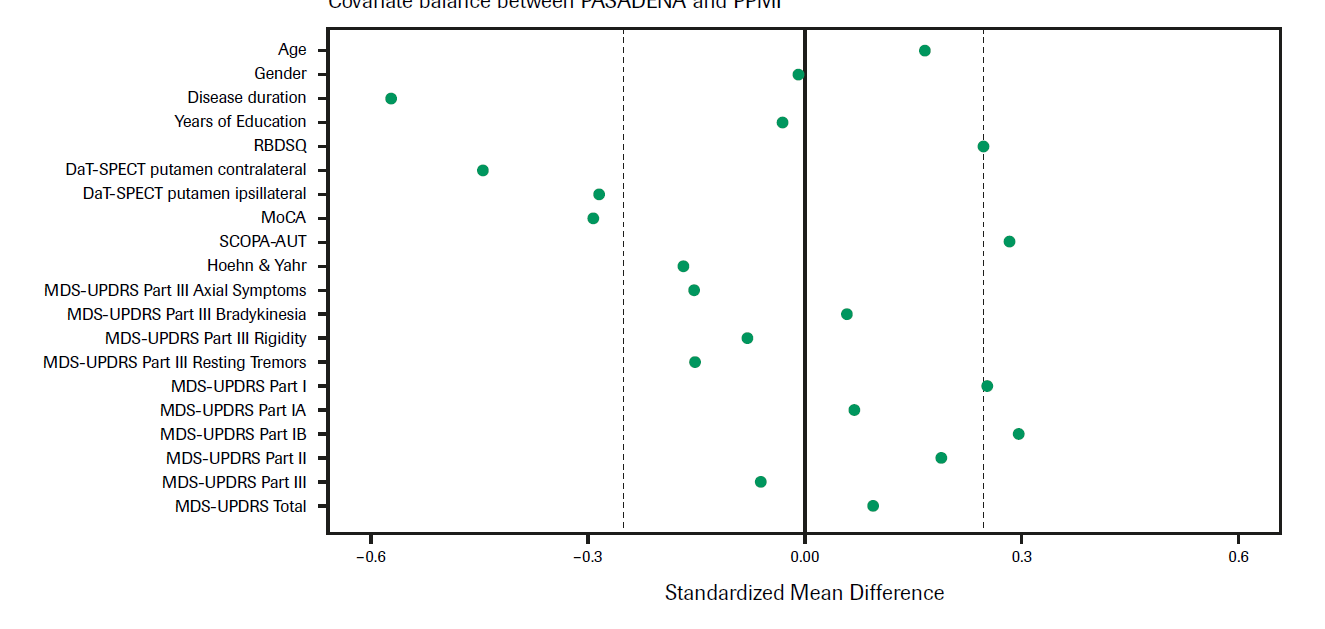

DaT-SPECT, dopamine transporter single-photon emission computerized tomography; MDS-UPDRS, Movement Disorder Society Unified Parkinson’s Disease Rating Scale; MoCA, Montreal Cognitive Assessment; PPMI, Parkinson’s Progression Marker Initiative; RBDSQ, Rapid Eye Movement Sleep Behavior Disorder Screening Questionnaire; SCOPA-AUT, Scales for outcomes in Parkinson’s disease autonomic dysfunction.
